# A model simulation study on effects of intervention measures in Wuhan COVID-19 epidemic

**DOI:** 10.1101/2020.02.14.20023168

**Authors:** Guopeng Zhou, Chunhua Chi

## Abstract

**Background:** In the beginning of January 2020, new unknown virus pneumonia cases started to emerge in local hospitals in Wuhan, China. This virus epidemic quickly became a public health emergency of international concern by the WHO. Enormous amount of medical supplies as well as healthcare personals from other provinces were mobilized to support Wuhan. This current work tent to help people understanding how infectious disease spread and the purpose and consequences of various efforts based on simulation model.

**Method:** a simulation model was created using known parameters. R0 set to 3 and mean incubation time to be 7.5days. the epidemic was divided to 3 periods. Simulation would run 50 times to mimic different patient0 status. Personal activity index was used to mimic different level of control measures. 141427709 simulated patients were created. Cumulation number of patients at the end of period 1 (day50) is 2868.7±1739.0. Total infected patients could be 913396.5 ± 559099.9 by the end of period 2 (day70) in free transmission state. And at day90, total patients number is 913396.5 ± 559099.9.

**Conclusion:** COVID-19 is a novel severe respiratory disease. This will put great burden on the shoulder of healthcare workers as well as on medical hardware and supplements. Current strict control measures help to contain disease from spreading. An early detecting, reporting and fast reacting system needs to be setup to prevent future unknown infectious disease.

---

In the beginning of January 2020, new unknown virus pneumonia cases started to emerge in local hospitals in Wuhan, China. transmissions between human were seen by the middle of January. And patient number increased sharply. In order to block the virus transmission from entire China and even worldwide. This virus epidemic quickly became a public health emergency of international concern by the World Health Organization.

Wuhan municipal government issued an official order to stop all transportation between Wuhan and the rest of China. Public transportations within Wuhan were also stopped. By 9th, February 2020, China central government mobilized enormous amount of medical supplies as well as healthcare personals from other provinces to support Wuhan. Till 11^th^ February,18700 healthcare personals belong to 154 medical teams had arrived in Wuhan and started to work with local hospitals. Wuhan government also declared regulations on residence building control, which include identity check at building entrance and strict infection control measures in buildings with suspect patients or identified patients.

With all these intervention measures undertaken, and maybe more on the way, proper tools are need anticipate possible effects on epidemic control and to provide reliable information to support future decisions. Simulation also can help people understand how infectious disease spread and how to understand the purpose and consequences of various efforts.

This current work was based on simulation model build on known transmission parameters from published research papers.

## Method

According to Qun Li et al. [1], 2019-nCoV’s average incubation time is 7.5 days (95% CI 5.3-19.0), 95% percentiles is 12.5 days. In early stage of the disease, patients’ number would double every 7.4 days. Basic reproduction number (R0) was presumed to be 2.2 (95% CI 1.4-3.9). In Qun’s paper R0’s 95% CI was between 1.4-3.9. R0=2, 3 or 4 were used in model preparation. And 2 and 4 were ruled out because of obvious devotion from open reported data. So R0 was set at 3 in this simulation.

Patient incubation time after been transmitted was assigned by python numpy random.normal function, with parameters set as: loc (mean value of random serie)=7.5, scale (standard deviation) = 3.4. Values lower than 0 are converted to positive values. In this study, simulation cases can only transmit virus to new patients on the last day of incubation to make calculation simple.

According to media report and Wuhan government’s announcement, the 2019-nCOV epidemic could be divided into 3 periods.

Period 1: free transmission period (day 1 to day 50)

Period 2: city-lock-down period (day 51 to day 70)

Period 3: enforced control period (day 71 to day 90)

The simulation would run like the diagram below.

**Figure 1.**
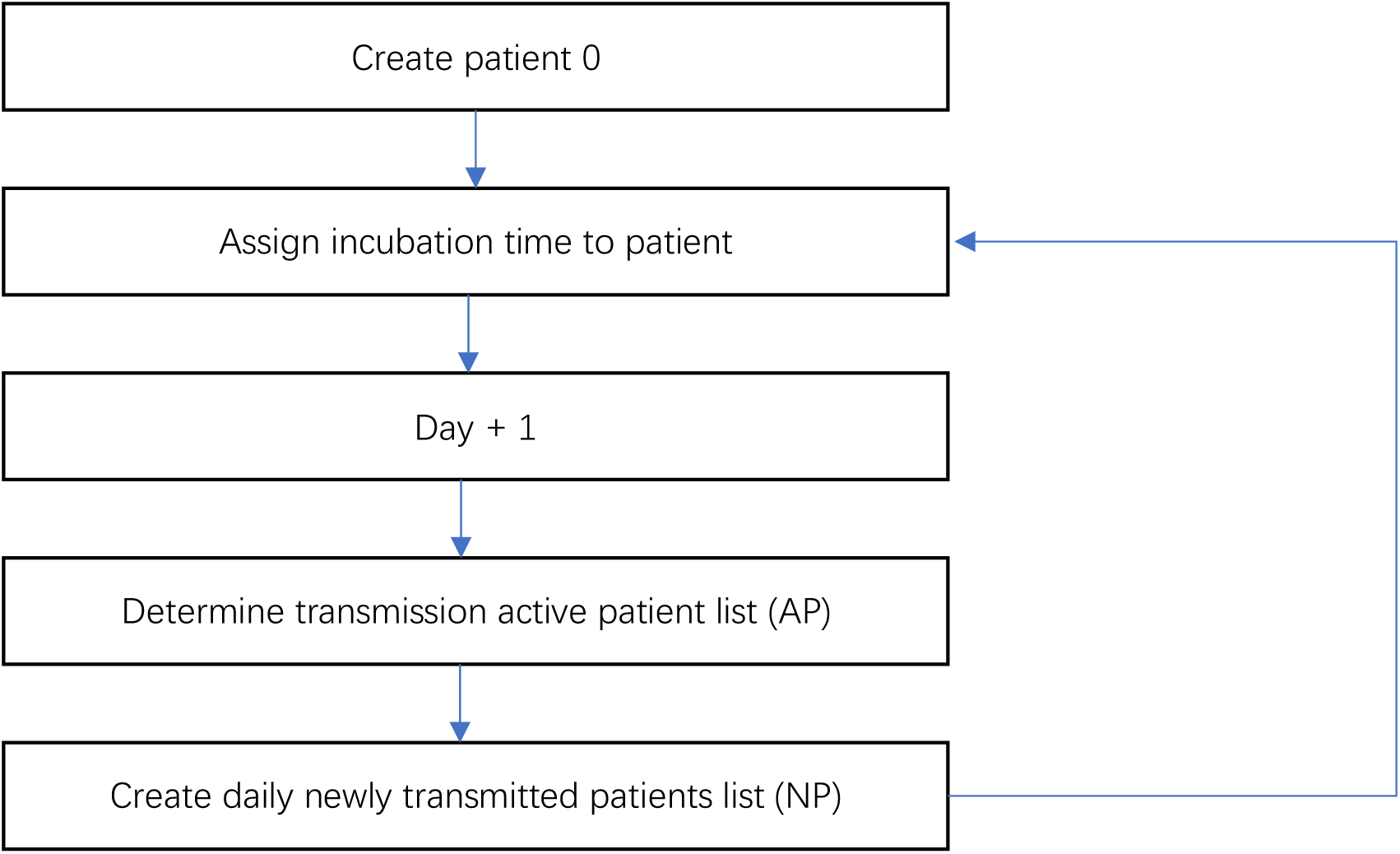
diagram of simulation process. NP = AP · R0 · PAI PAI: personal activity index, an artificial index created to demonstrate patients’ mobility status in this study. PAI=1 means no restriction of social activity, while PAI=0 means totally grounded (include in house isolation and all sorts of hospitalization).

Based on these parameters a simple simulation model was accomplished with python 3.6. All calculation was run on a mac pro notebook with 3.1 GHz duel core CPU and 8GB memory. Simulation would run 50 times to mimic different patient0 status.

Data were shown as mean ± standard deviation.

## Results

Simulations were run by 50 times. 141427709 simulated patients were created with incubation time 7.5 ± 3.4 days. The distribution of incubation time are show in Figure 2.

**Figure 2.**
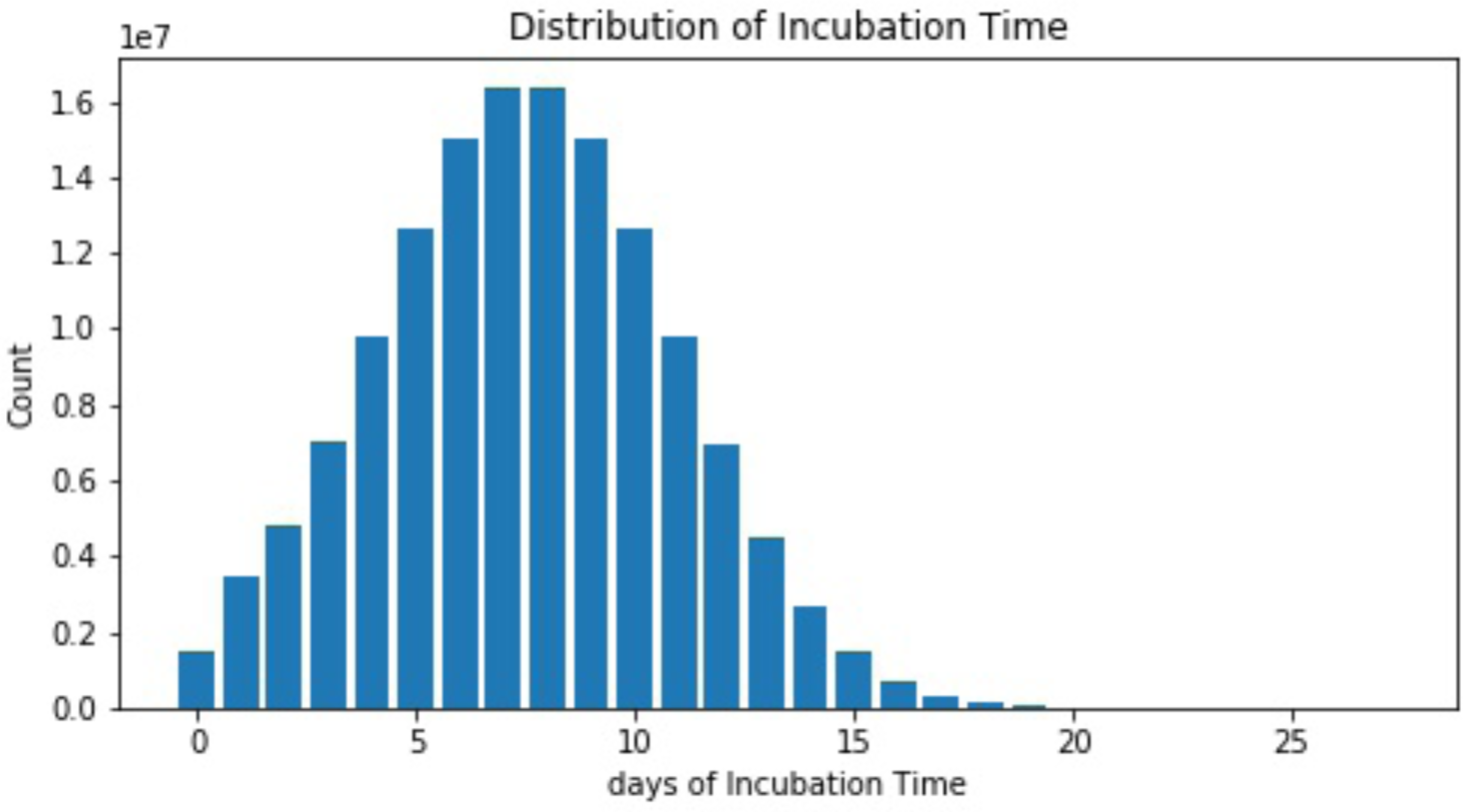
distribution of patient incubation time.

By simulation, Cumulation number of patients at the end of period 1 (day50) is 2868.7±1739.0. Total infected patients could be 913396.5 ± 559099.9 by the end of period 2 (day70) in free transmission state. And at day90, total patients reached a horrible number: 913396.5 ± 559099.9 (Table 1). And among these patients, 5% (more than 45000 patients) would be critically ill [2].

**Table 1.** patient number estimation under free transmission

| date | Cumulation number of<br>patients | Daily transmission<br>active patients | Daily new patients |
| --- | --- | --- | --- |
| Day 50 | 2868.7±1739.0 | 141.4±86.8 | 424.2±260.3 |
| Day 70 | 52185.4±31621.4 | 2582.9±1554.6 | 6973.5±4197.5 |
| Day 90 | 913396.5±559099.9 | 45006.34±27555.4 | 121516.7±74399.7 |

According to simulation data, cumulative amount of infected patients (Figure 3), daily transmission active patients (Figure 4), and daily new patients number (Figure 5) would keep raising if PAI was above 0.3 which might cause a prolonged epidemic of a dreadful virus respiratory disease. When PAI was lower or equal to 0.3, a plateau would emerge. With stricter measures taken (PAI<0.3), the daily new patients number started to decline in period 3 respectively, see in Figure 6 to Figure 15.

**Figure 3.**
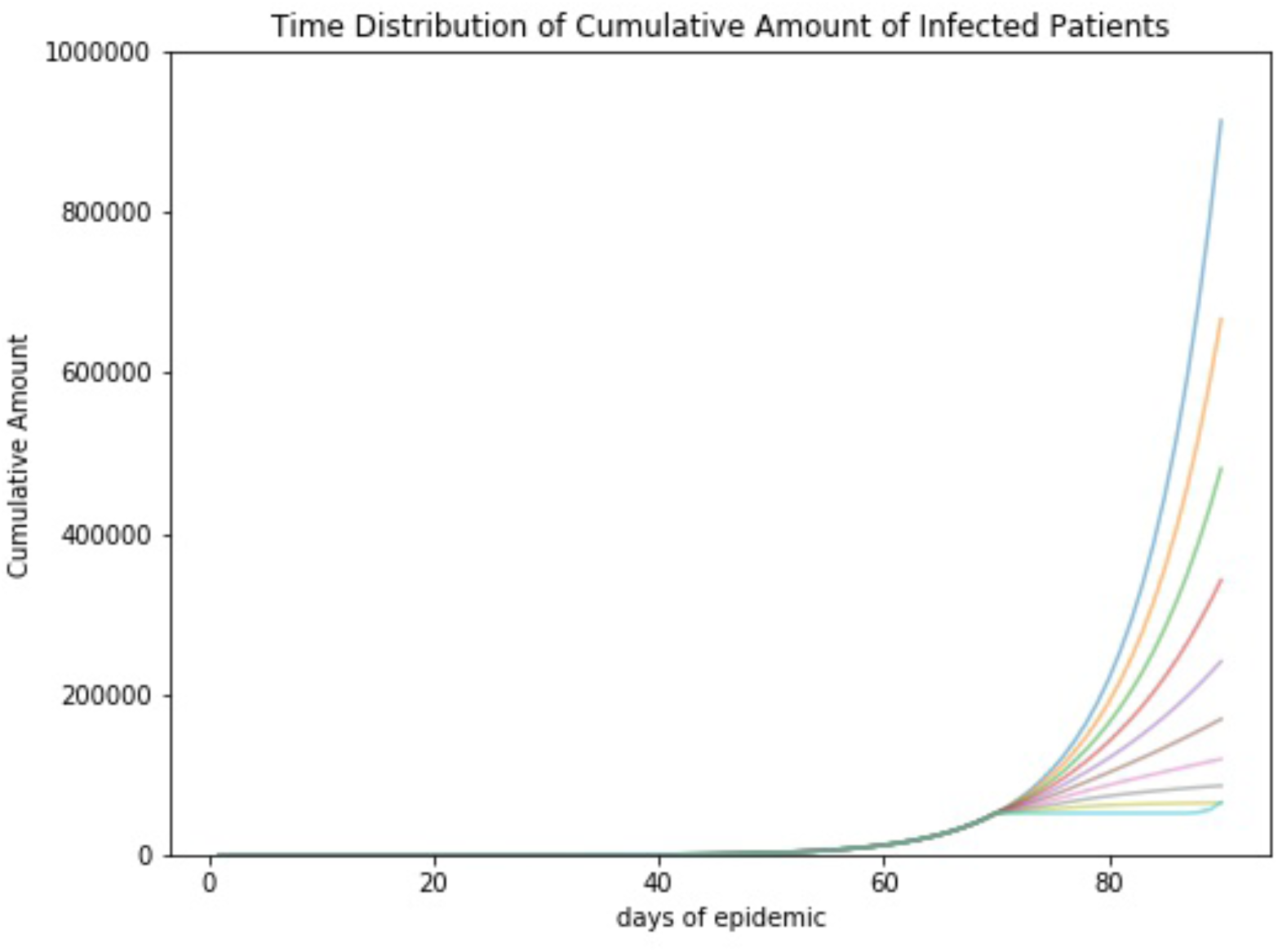
time distribution of cumulative amount of infected patients (curves from top to bottom: PAI = 0.9, 0.8, 0.7, 0.6, 0.5, 0.4, 0.3, 0.2, 0.1, 0 respectively)

**Figure 4.**
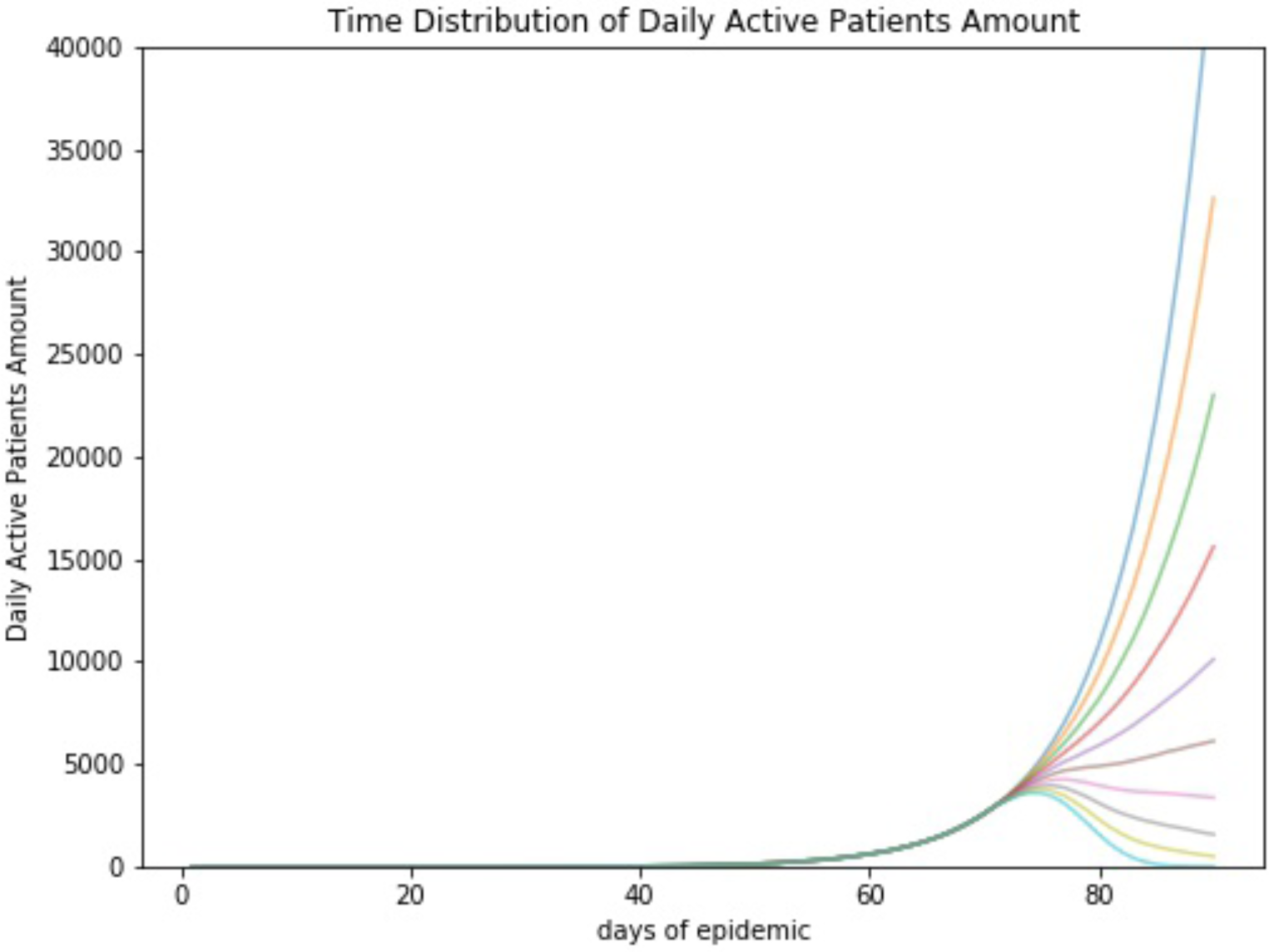
time distribution of daily transmission active patients amount (curves from top to bottom: PAI = 0.9, 0.8, 0.7, 0.6, 0.5, 0.4, 0.3, 0.2, 0.1, 0 respectively)

**Figure 5.**
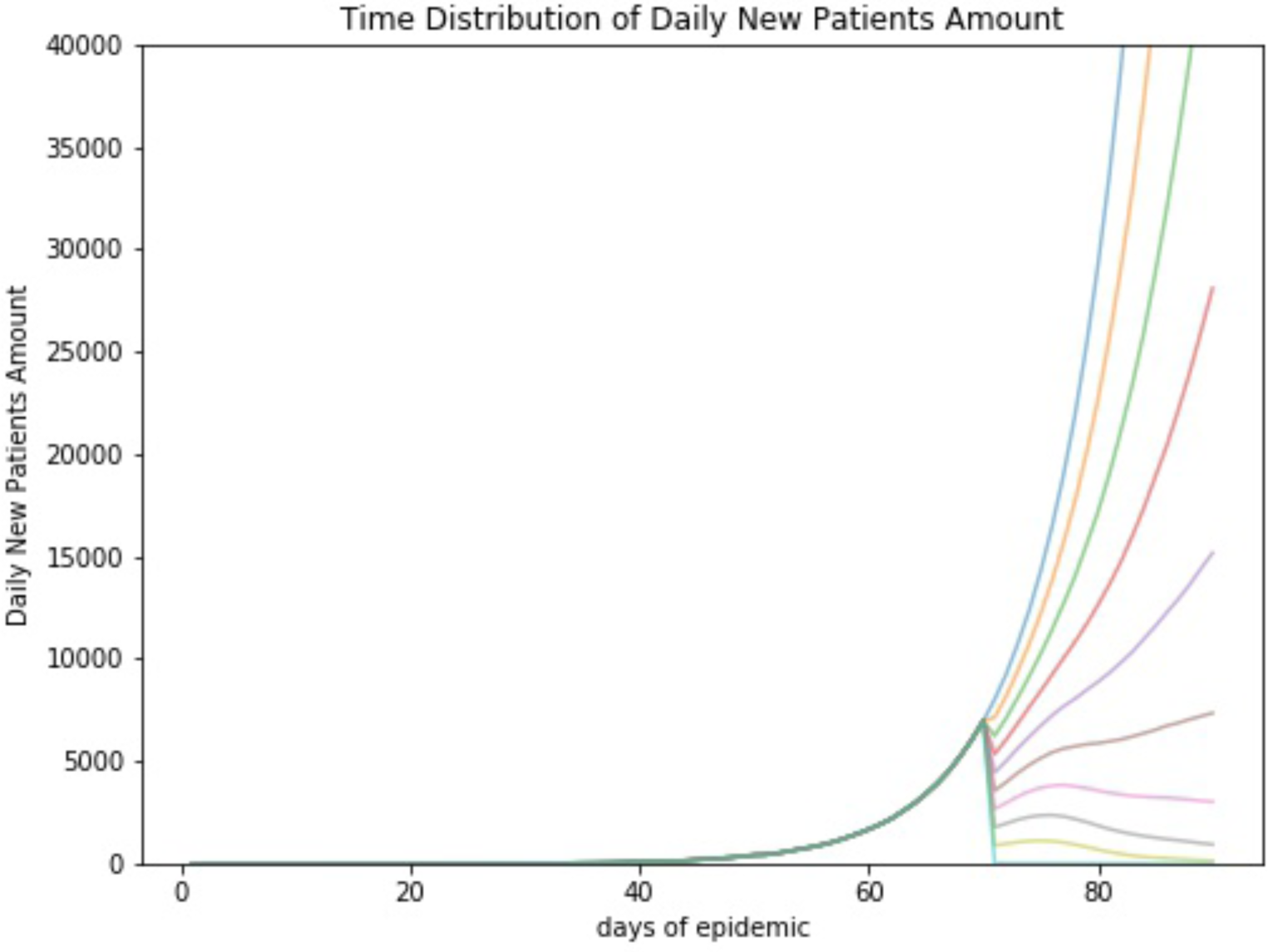
time distribution of daily new patients amount (curves from top to bottom: PAI = 0.9, 0.8, 0.7, 0.6, 0.5, 0.4, 0.3, 0.2, 0.1, 0 respectively)

**Figure 6.**
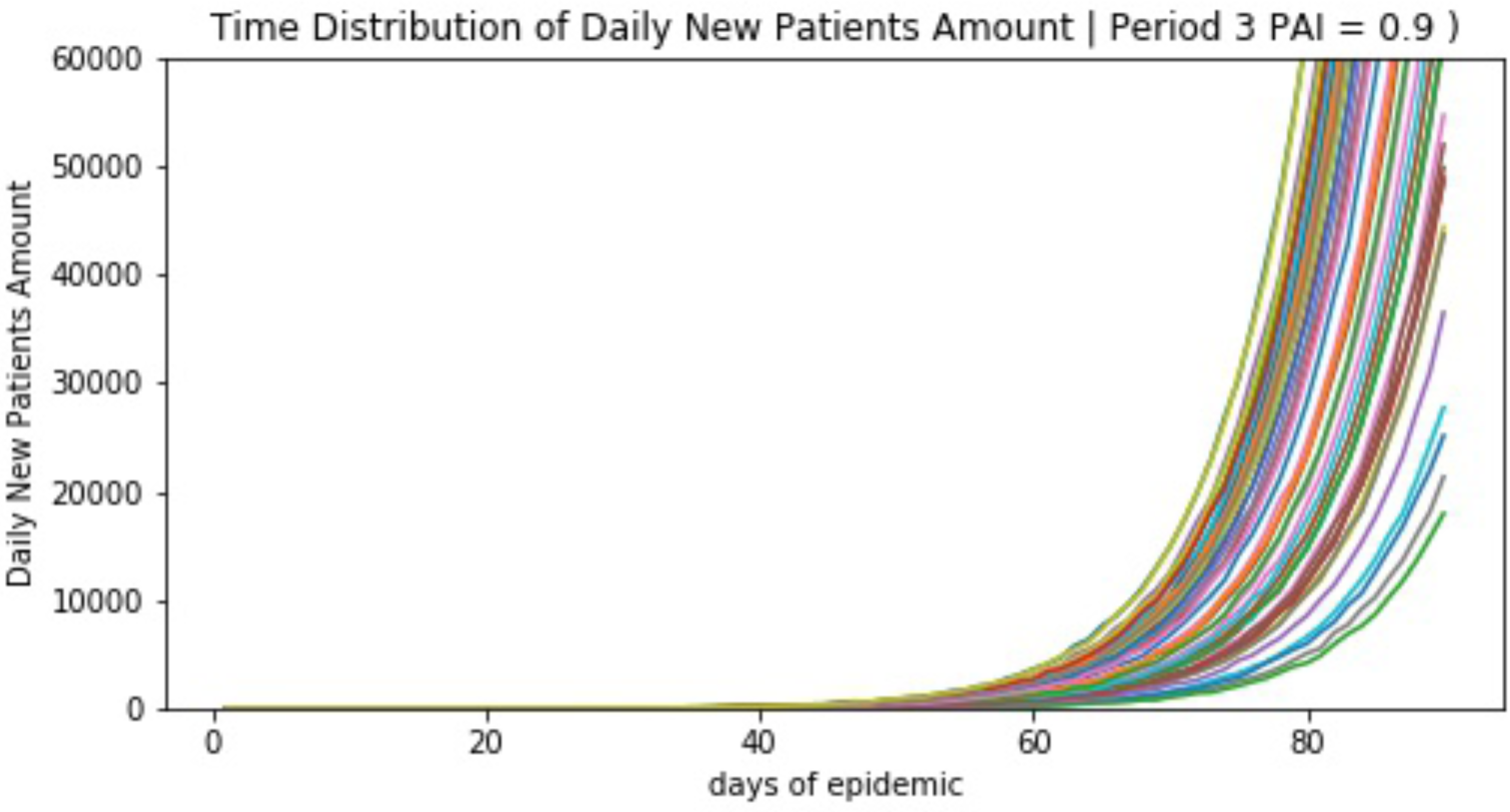
simulated time distribution of daily new patients in period 3 (PAI=0.9)

**Figure 7.**
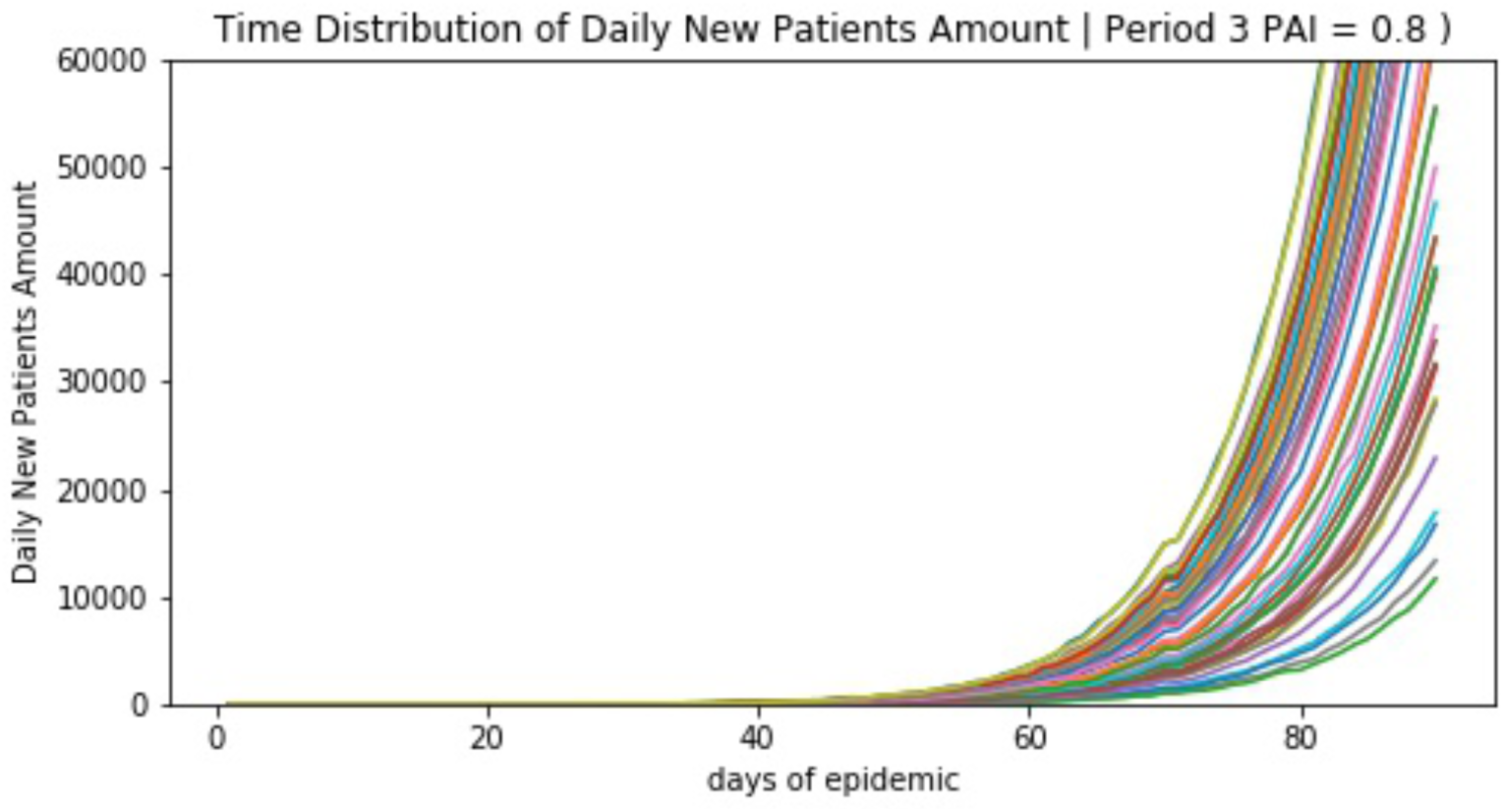
simulated time distribution of daily new patients in period 3 (PAI=0.8)

**Figure 8.**
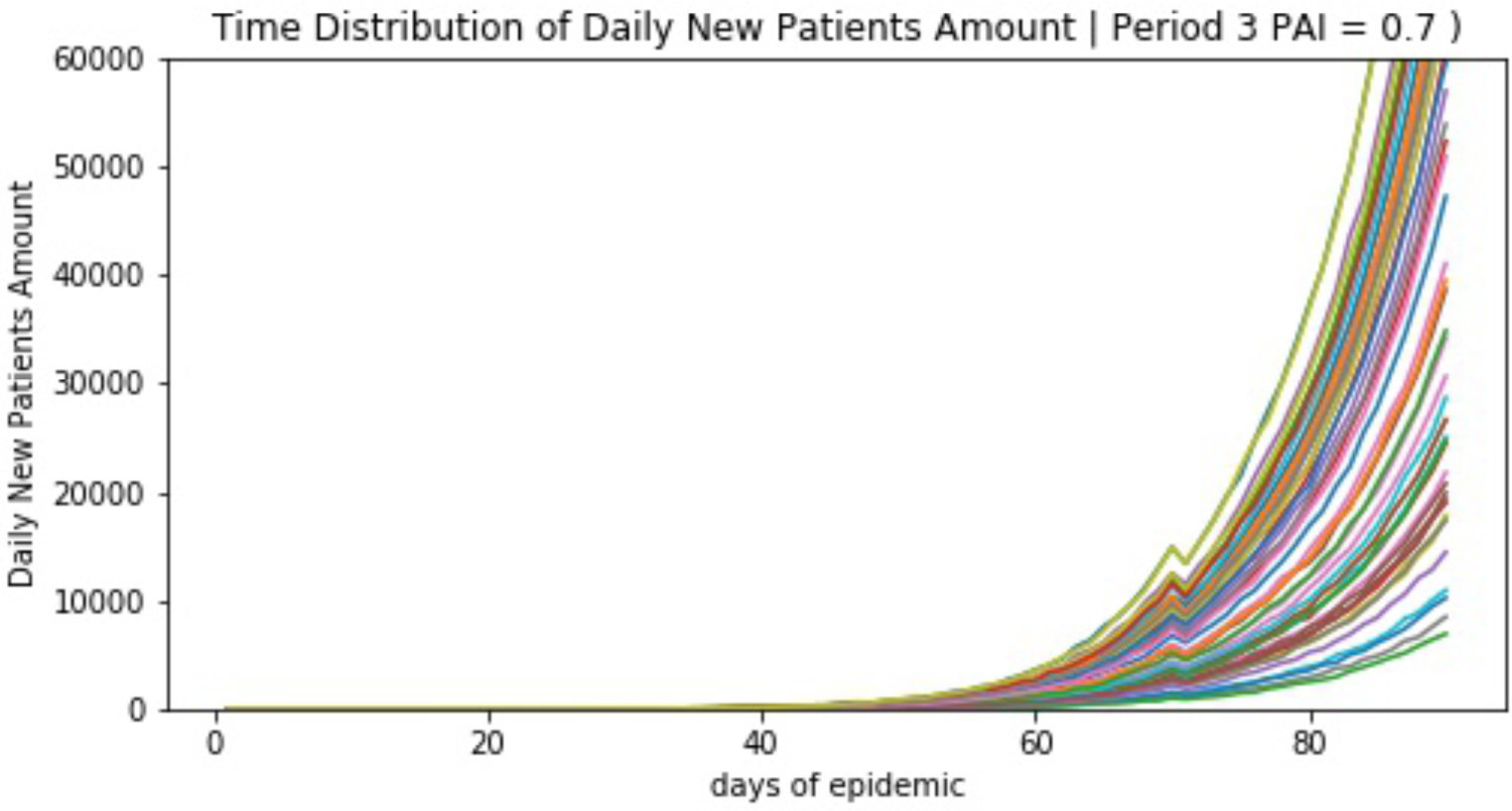
simulated time distribution of daily new patients in period 3 (PAI=0.7)

**Figure 9.**
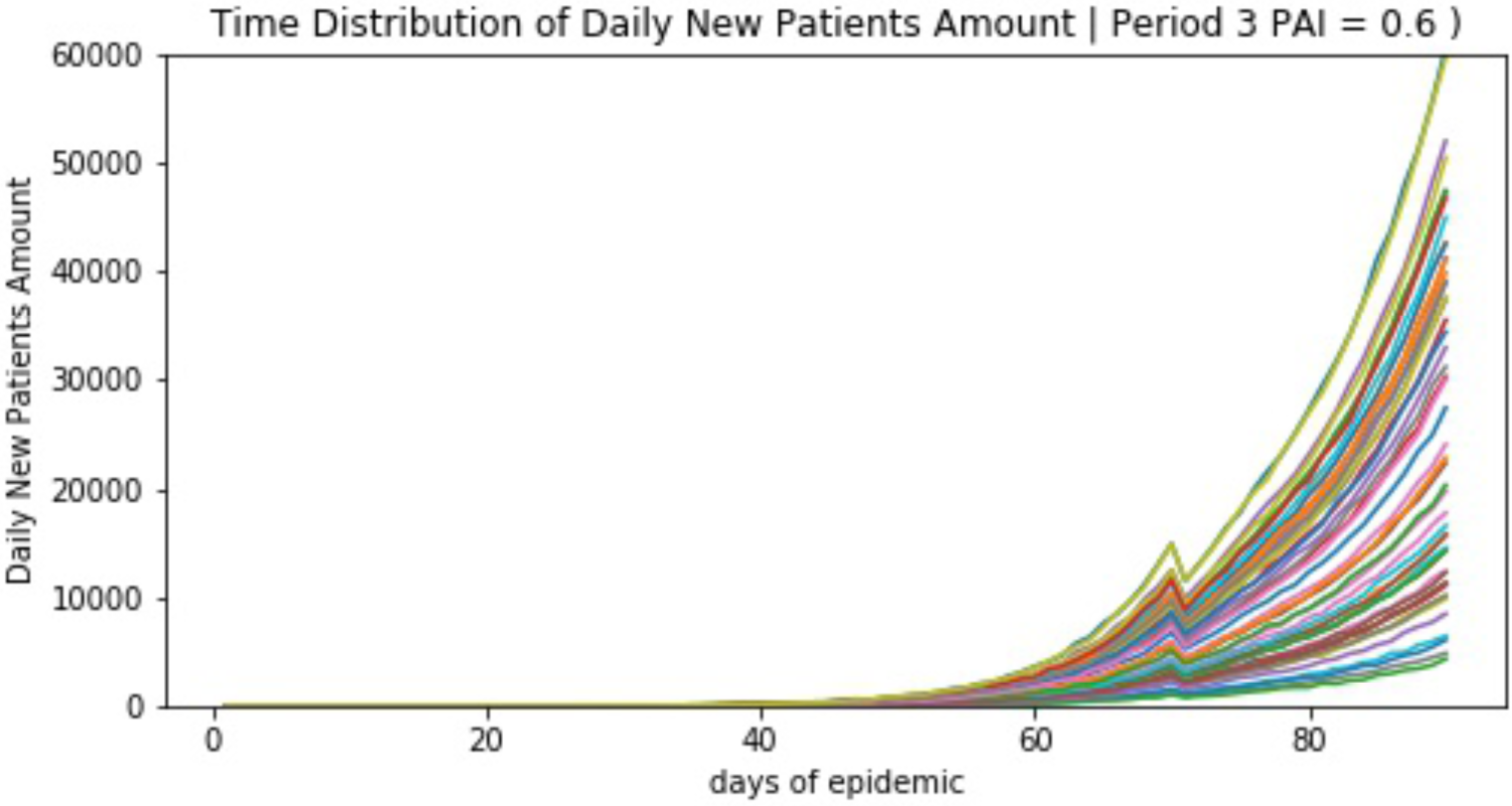
simulated time distribution of daily new patients in period 3 (PAI=0.6)

**Figure 10.**
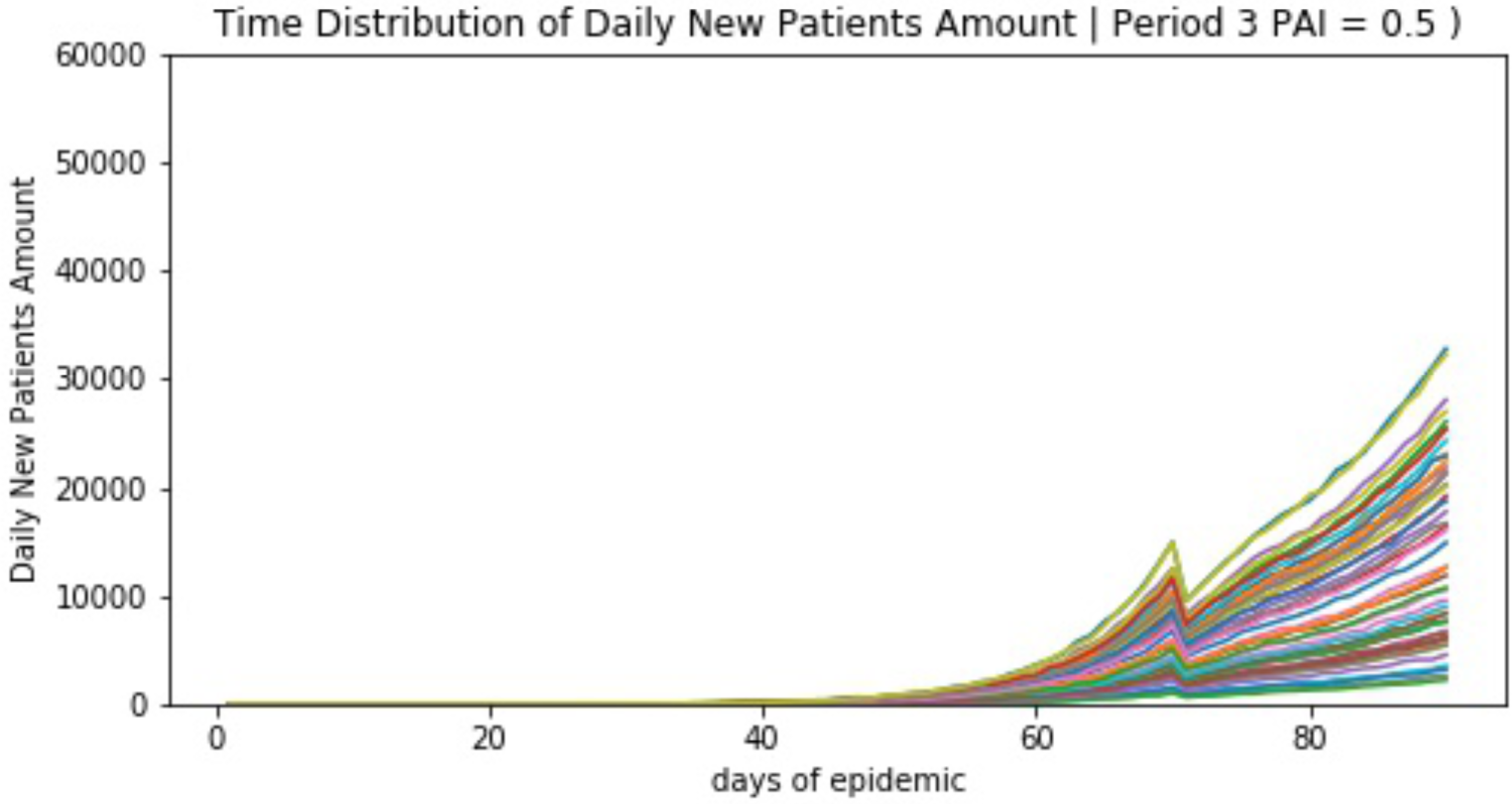
simulated time distribution of daily new patients in period 3 (PAI=0.5)

**Figure 11.**
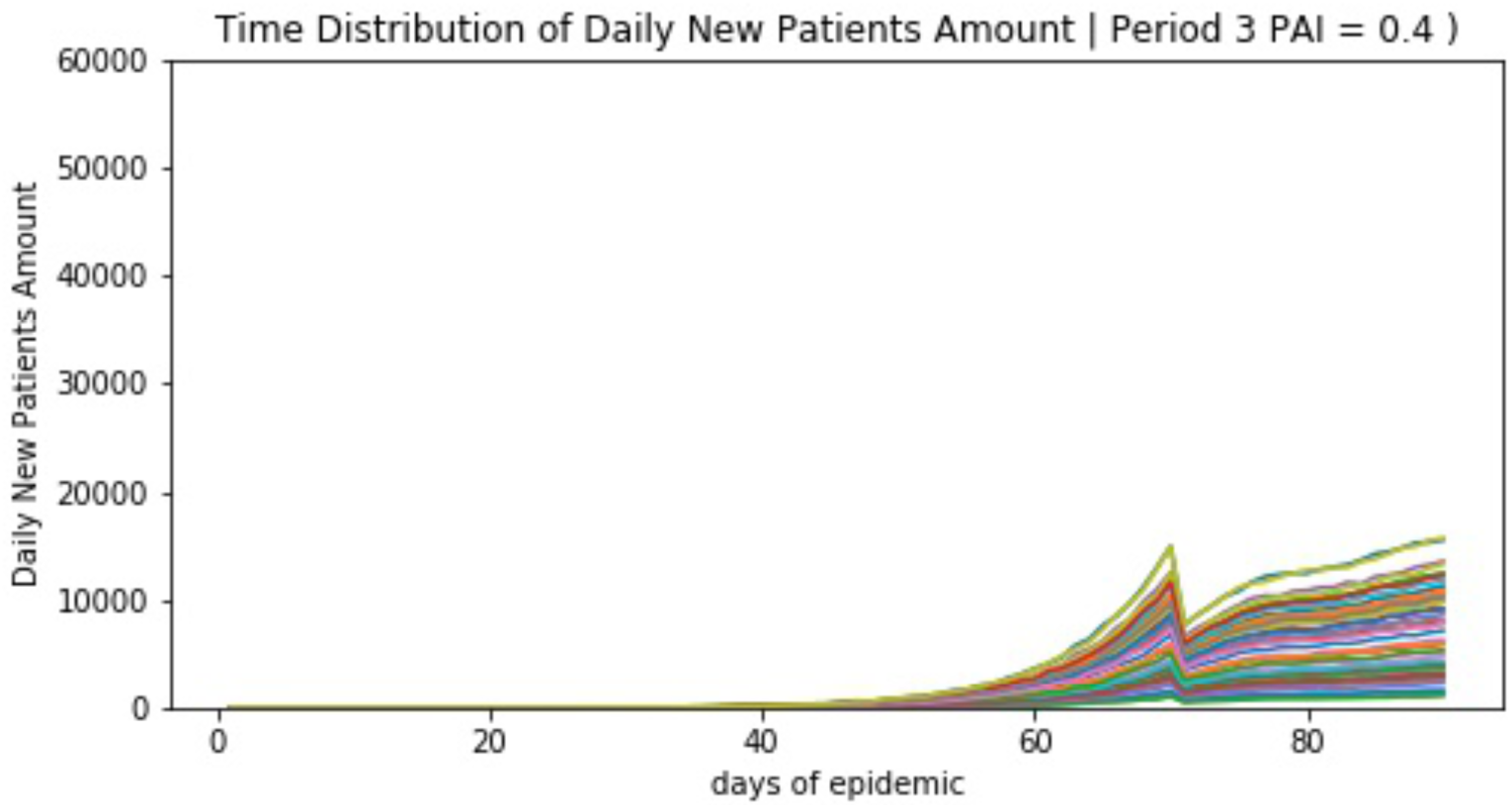
simulated time distribution of daily new patients in period 3 (PAI=0.4)

**Figure 12.**
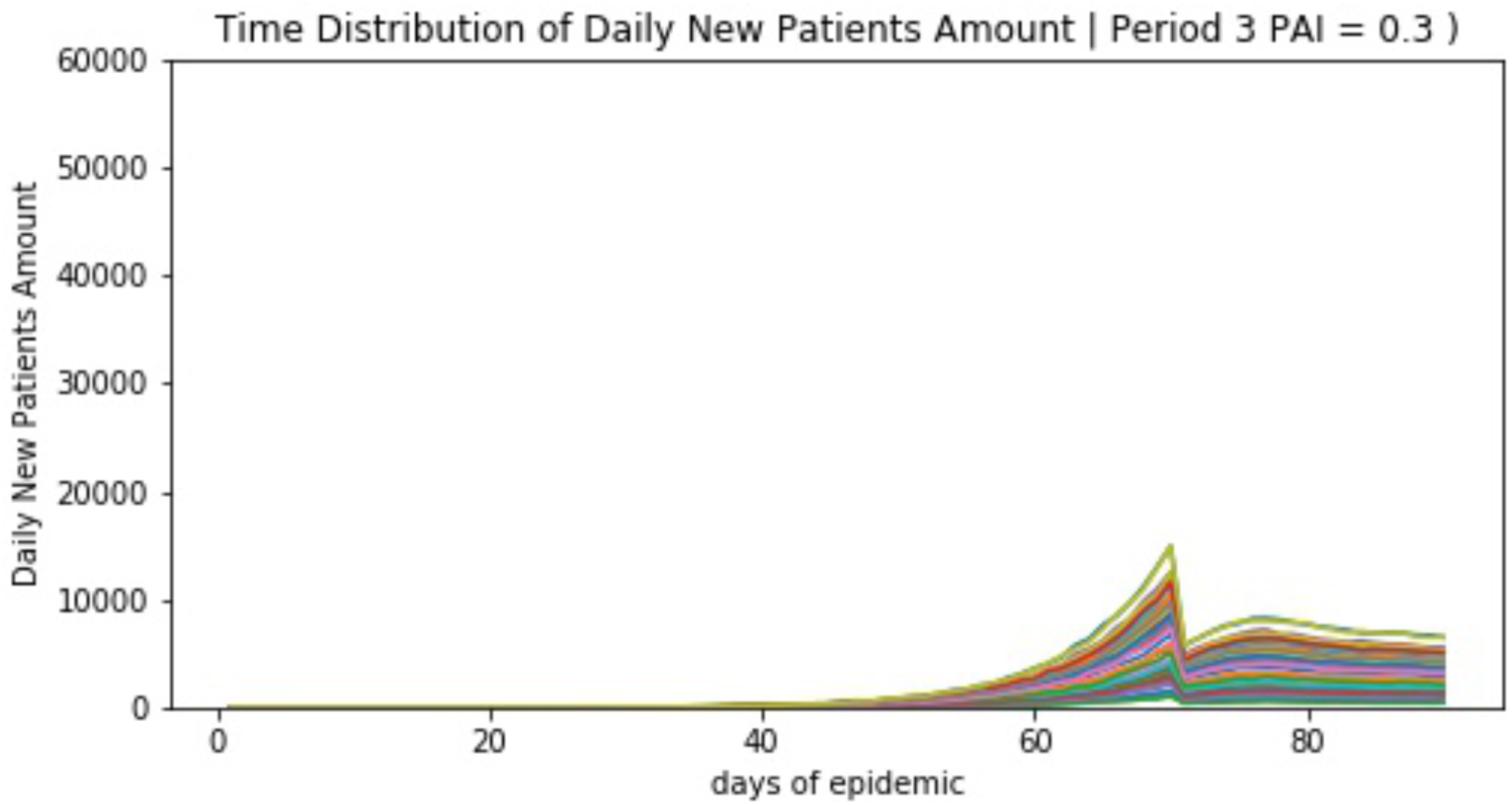
simulated time distribution of daily new patients in period 3 (PAI=0.3)

**Figure 13.**
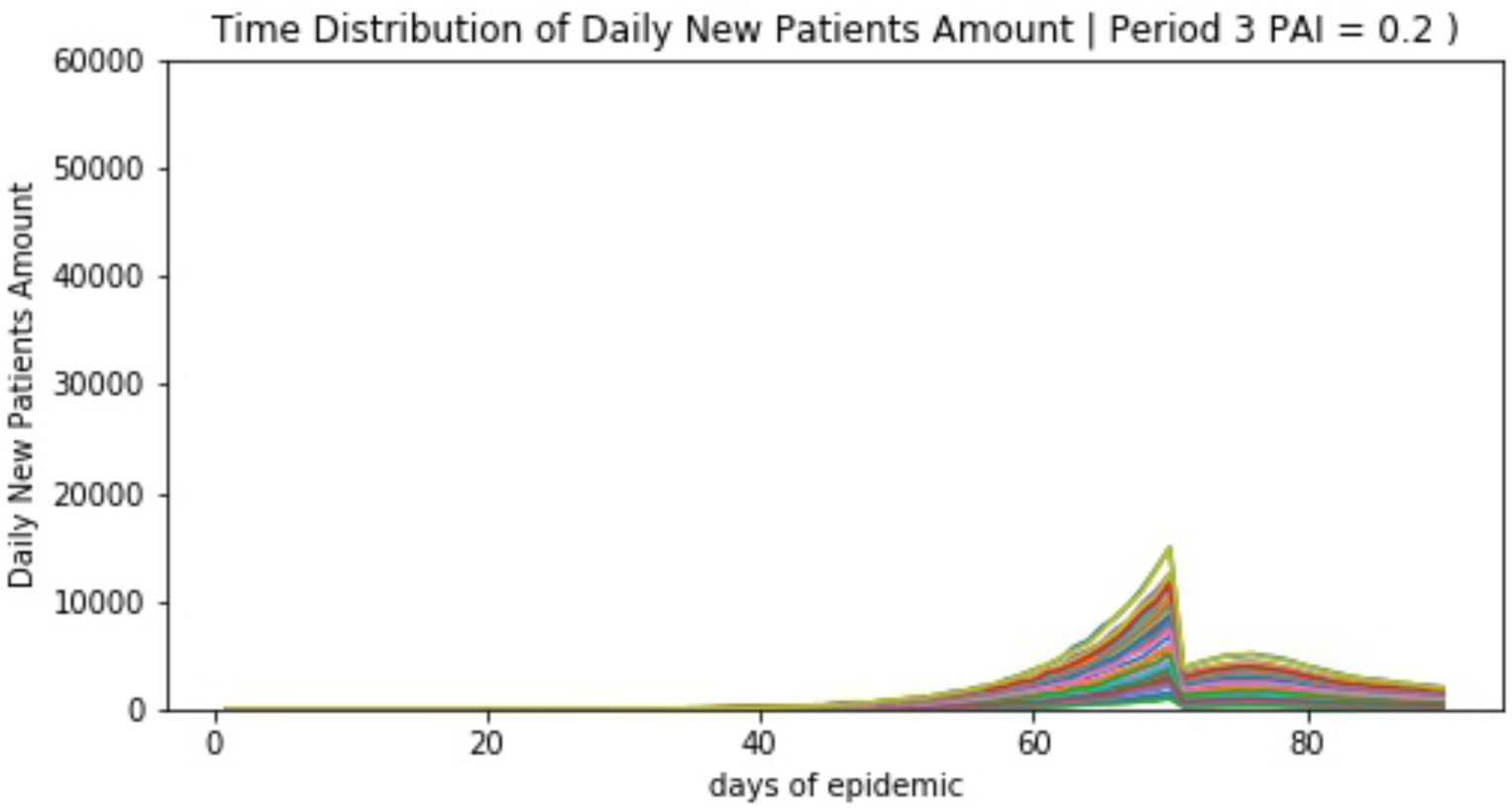
simulated time distribution of daily new patients in period 3 (PAI=0.2)

**Figure 14.**
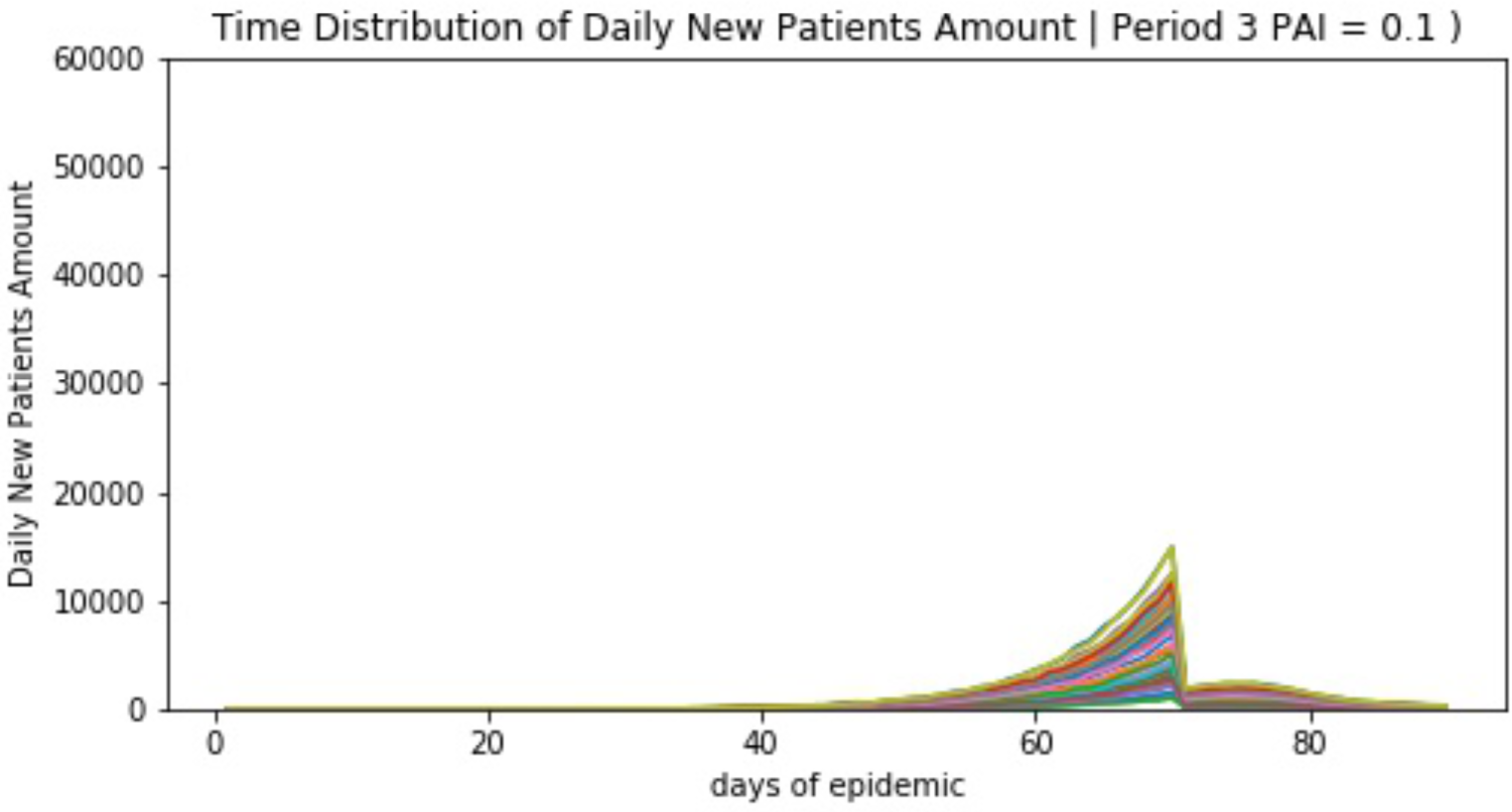
simulated time distribution of daily new patients in period 3 (PAI=0.1)

**Figure 15.**
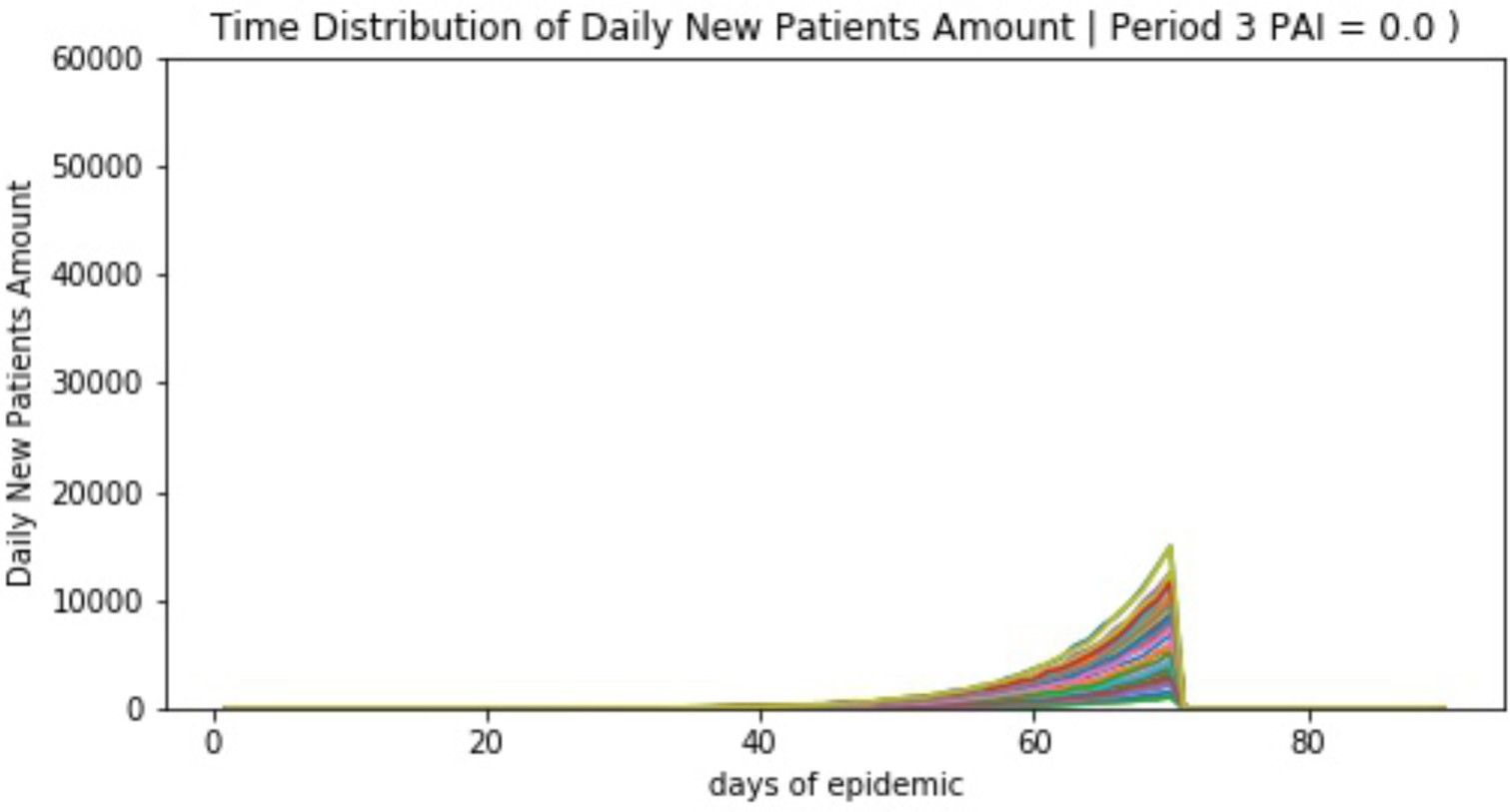
simulated time distribution of daily new patients in period 3 (PAI=0.0)

## Discussion

4 unknown pneumonia patients were spotted by 29^th^ December 2019 in Wuhan China. all patients had history related to Huanan Seafood Market. Similar cases kept emerging. Qun Li et al. collected 435 cases by the middle of January 2020[1]. Average incubation time assumed to be 7.5 ± 3.4 days (95% CI 5.3-19.0). In the early stage of disease, patients number would be doubled every 7. 4days.R0 was calculated to be 2.2 (95% CI 1.4-3.9).

National Health Commission issued an order to treat 2019-ConV as a category B infectious disease, controlled as category A (same policy as SARS). On the day of 23 January 2020, Wuhan municipal government decided to lock down the city. All public transportation except emergence services between this giant south China city and the rest of China were stopped. In-city public transportation services were also on held. China started to mobilize enormous medical resources include healthcare teams to help Wuhan.

Under multiple measures meant to contaminate this novel virus, diagnosed and suspected cases kept increasing. Among the infected patients, 5% or more were in critical condition and were located in intensive care units. When could the public see ending of this epidemic become a common concern. We used computer model simulation to show effects of different level of efforts in containing this virus from spreading. To make things simple, restrictions of patients’ activities (including in-hospital treatment, cabin hospital stay, and in-house surveillance) were represented as personal activity index PAI.

Read, Jonathan’s preprint work showed a R0 to be 3.6-4.0[3]. In our simulation study, data show simulated patients to be 2868.7±1739.0 by the end of period 1 (day50). Data from China Disease Control and Prevention Center shows cumulated patients number was 31728, daily new patients number was 3911 by 10^th^ February, 2020 (http://2019ncov.chinacdc.cn/2019-nCoV/). This information confines within simulated data of our model if we set PAI of period 2 to be 0.9. When it came to end of period 2, simulated patients’ number had reached 52185.4±31621.4. We can deduce that city lock-down did not work as supposed.

Till 11^th^ February 2020, more than 154 medical teams, 18700 healthcare personels were sent to Wuhan. A whole population body temperature monitoring program was also undertaken to identify fever patients. And local CDC announced to stop using time consuming RNA test as common diagnosis standard. Local residencies were locked down to minimize inter people transmission.

How these measures would affect disease spreading is still not knowing. So we simulated the effects by set PAI from 0.9 to 0. PAI = 0.9 means no change in current people behavior while PAI = 0 means total grounded (include hospital stay and in-house surveillance).

With simulated data, we found that:

Wuhan have 81700 hospital beds in nearly 300 hospitals among which 61 top tier hospitals in a 2018 government report (http://wjw.wuhan.gov.cn/upload/file/20191205/1575536693972018707.pdf). In free transmission stage, cumulated patients number reached 52185.4±31621.4 on day 70. Critical care rate is 5% according to Zhong’s preprint manuscript [2]. So more than 2600 patients needed intensive care. This huge infectious patient number would stop all Wuhan ICUs from proper function.

At the beginning of stage 3, different PAI were used to calculate future trends in disease epidemic. We found that new daily patients number seem to stay in plateau when PAI =0.3. When PAI reduced to 0.2, a decline in daily new patient could be seen. If PAI could be restricted to 0.1, an ideal state of zero new daily patient would show up at the end of period 3.

## Conclusion

1. COVID-19 is a novel severe respiratory disease. It can cause strong inter people transmission. Enormous difficulties could be anticipated if not contained in early stage.
2. 5% of COVID-19 patients need intensive care. This will put great burden on the shoulder of healthcare workers as well as on medical hardware and supplements.
3. Current strict control measures help to contain disease from spreading. And long-term effect relates to how well Wuhan fulfill this task.
4. An early detecting, reporting and fast reacting system needs to be setup to prevent future unknown infectious disease.

A respiratory disease spreading involves numerous factors. This simulation model uses only arbitrary data to make the calculation. And also, parameters are simplified to make the calculation faster, the results may be varying from real data. The model should be modified using actual data in order to get more accurate results.

## Data Availability

no actual patient data were used in this research

## Notes

### Competing Interest Statement

The authors have declared no competing interest.

### Funding Statement

no funding

